# A modified SEIR Model with Confinement and Lockdown of COVID-19 for Costa Rica

**DOI:** 10.1101/2020.05.19.20106492

**Authors:** Tomas de-Camino-Beck

## Abstract

The fast moving post-modern society allows for individuals to move fast in and between different countries, making it a perfect situation for the spread of emerging diseases. COVID-19 emerged with properties of a highly contagious disease, that has spread rapidly around the world. SIR/SEIR models are generally used to explain the dynamics of epidemics, however Coronavirus has shown dynamics with constant non-pharmaceutical interventions, making it difficult to model with these simple models. We extend an SEIR model to include a confinement compartment (SEICR) and use this to explain data from COVID-19 epidemic in Costa Rica. Then we discuss possible second wave of infection by adding a time varying function in the model to simulate cyclic interventions.

## 1 Introduction

The fast moving post-modern society allows for individuals to move fast in and between different countries, making it a perfect situation for the spread of emerging diseases. COVID-19 has emerged with the properties of a very aggressive infectious disease with several features that allows for fast spread [1], giving rise to a pandemic that started in China and has spread fast around the world. Since there is no vaccine for COVID-19, all control strategies have focused on non-pharmaceutical interventions (NPI) to contain and slow down the spread of the disease and “flatten the curve” [8].

Because of the spatio-temporal scale of the spread of COVID-19, modelling approaches have to be used to understand COVID-19 dynamics. Epidemiological models have shown to be very valuable for designing such control strategies. Although they miss some of the details of the dynamics, and assume random mixing, some general control strategies,like social distancing, confinement, quarantine and others, can be studied from analysis of these systems of equations, and they have proven effective even under lack of data [6], since they can capture some of the fundamental aspects of disease dynamics, they can be used to create possible epidemic scenarios.

Several SIR models have been applied to Coronavirus for different regions [5, 2, 10, 3], and most of the focus has been the estimation of *R*_0_, the basic reproductive number, predictions on the number of infected and mortality, and comparisons between regions. Additionally, SIR models can be used to test hypothesis on the dynamics of epidemics by changing the model structure, incorporating compartments and/or including time varying parameters, these models can then be adjusted and validated with data, and then used for forecast and establishment of possible scenarios.

In Costa Rica, the increase in number of cases showed sub-exponential growth since the onset of the epidemic, with low number of cases and mortality, and reportedly no community infections. The reason is attributed to government early country wide NPI that have been considered to be effective [12, 11]. Based on a goverment public report [4], a second wave its expected, but no mathematical model has been provided to create possible scenarios, and there is uncertainty on how relaxation of NPI measures might have an effect.

In this brief paper, we introduce an SEIR based model to account for the dynamics introduced by confinement. Adding confinement we believe we can introduce discrete external forces like social distancing and lock downs, that change the dynamics by dynamically changing the availability of susceptible people in the system. Based on this model it is possible to create cyclic enforcement-relaxation of social distancing measures to forecast the impact of a possible second wave. We use the model to study COVID epidemic in Costa Rica.

## 2 Model Description

We use an extension of an SEIR model to incorporate confinement. As with all SEIR models, we define *S* to be susceptible, *E* exposed, *I* infected and *R* recovered (or dead). We introduce compartment *C* for confined individuals, that is, individuals whose movement are restricted and effectively removed from the susceptible population by strong NPI, like lock downs, closure of retail and entertainment, parks, and vehicular restrictions. In figure 1a we show our SEICR model.

**Figure 1:**
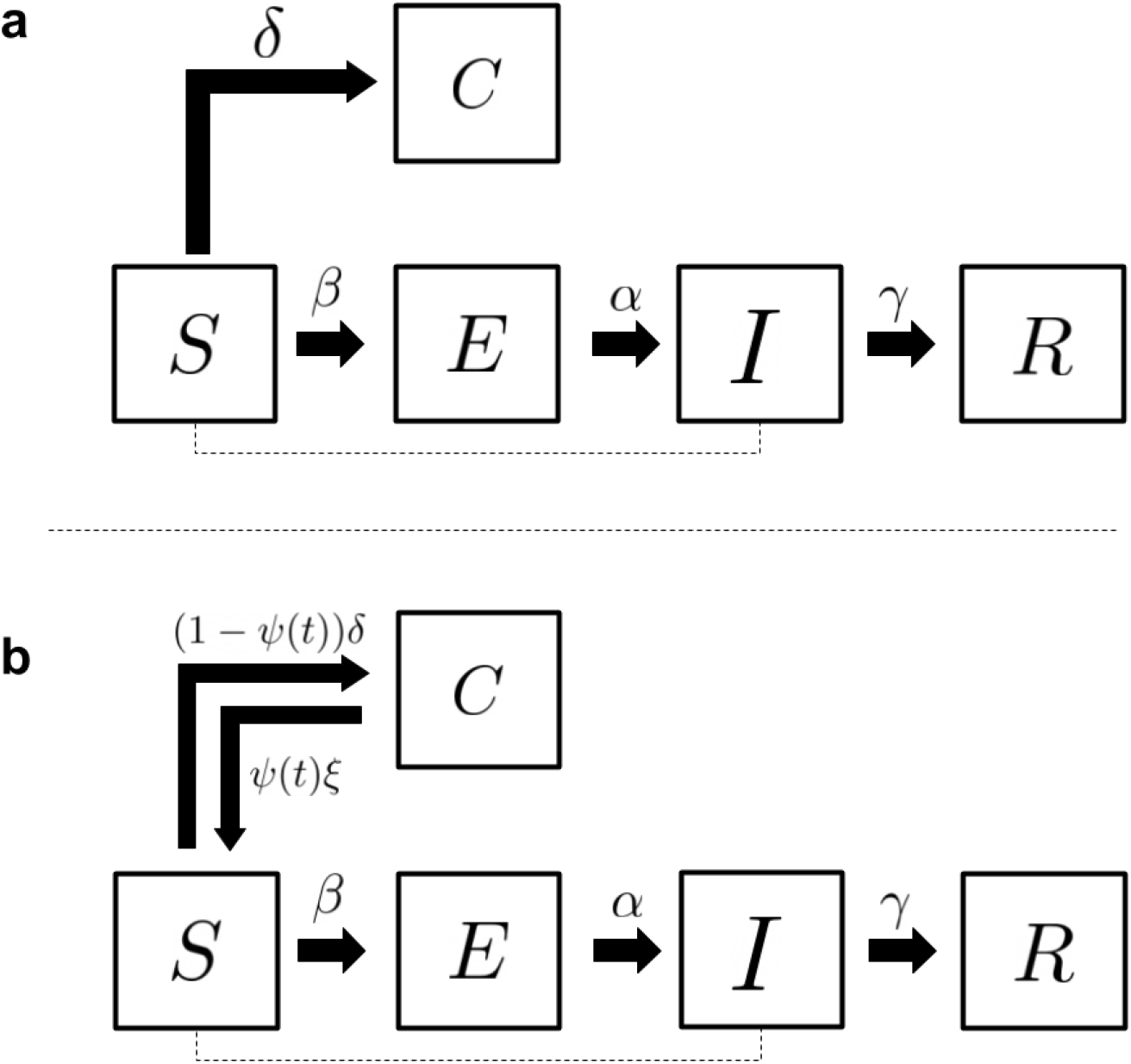
a. SEICR compartment model, b. SEICR model with added movement from confinement to susceptible

The parameters are described in Table 1. This model is a standard SEIR model, where there is contact between susceptible and infected individuals, described by a mass action term with infection rate *β*. There is an incubation period given by *E* moving to infected at rate *α*, and then infected *I* move to the removed compartment *R* with rate *γ*, where we include recovered and dead individuals. The model does not include any type of migrations or natural birth and death.

**Table 1:** COVID-19 SEICR Parameters

| Parameter | Value | Description | Observation |
| --- | --- | --- | --- |
| $\beta$ | ? | Transmission rate | To be estimated |
| $\alpha$ | 1/6 | Incubation rate | Based on CR reports |
| $\delta$ | ? | Confinement rate | To be estimated |
| $\gamma$ | 1/21 | Removed rate | Based on CR reports |

By including compartment *C*, susceptible individuals in the population can move to confinement at rate *δ* and stay there, reducing the number of available people that can get infected in the dynamics. The full system is shown in equation 1. A Similar model have been developed in [9] by allowing movement of susceptible and infected to a quarantine compartment, in our model we only Allow movement of susceptible.

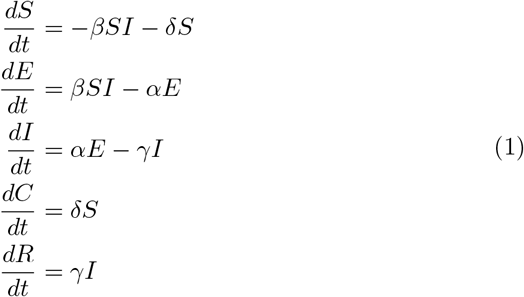

## 3 Modified SEICR

This second model is the same as described in equation 1, however we include a return to susceptible with rate *ξ*, and a discrete time variant function *ψ*(*t*) ∊ {0, 1} (see figure 1b), that takes the value 0 when measures of lock down are applied and 1 when the measures are relaxed with rate *ξ* ≤ *δ*. The full system is shown in equation 2

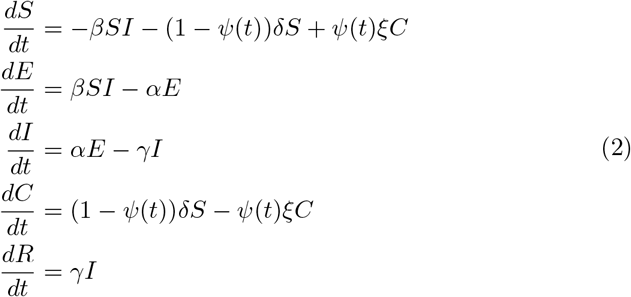

Note in equation 2 how the movement between susceptible and confinement compartments occur only in one direction. If *ψ*(*t*) = 0 then there is movement from susceptible to confinement, and in the opposite direction if *ψ*(*t*) = 1. Note also that, when *ψ*(*t*) = 0, the system reduces to that of equation 1.

## 4 Methodology

With data available of number of active COVID-19 cases for Costa Rica, we use data from the beginning of the infection on February 5 until May 3rd, where some NPI were relaxed, to fit our SEICR model. Then we used the full data set from February 3 to May 14, to study possible new outbreaks with model SEICRS.

We first tried to fit the model described in equation 1, to estimate parameters *β* and *δ*, and using parameter values as described in table 1. We forced the fitting with incubation period *α* = 1/6 and *γ* = 1/21. We first used differential evolution to reduce parameter search space, and later we used non-linear fitting with gradient descent.

To fit the system of equations, we created a parametric function to solve numerically with Runge-Kutta methods for the system, using an initial population 5 million susceptible individuals and starting with only one infected individual, and no individuals in any other compartment.

Once we found parameter values for *β* and *δ* for the SEICR model, we extend the model to include the return to susceptible class, as described by the SEICRS model, to then create possible scenarios with cyclcic NPI.

For all calculations, non-linear fitting and numerical solutions, we use the software *Mathematica* 12.

## 5 Results

### 5.1 Model Fitting

We adjusted the number of active cases to the system in equation 1, given fixed parameters for *α* and *γ* from table 1. The adjusted values for *β* and *δ* are given in table 2. The model with best best fit parameters, 99% confidence intervals, and an *R*^2^ = 0.99 is shown in figure 2

**Table 2:**
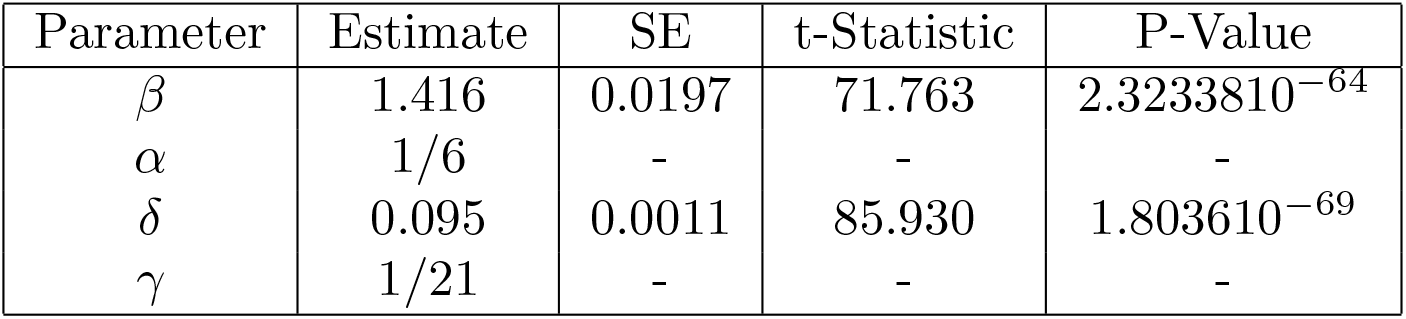
Adjusted SEICR Parameters

| Parameter | Estimate | SE | t-Statistic | P-Value |
| --- | --- | --- | --- | --- |
| $\beta$ | 1.416 | 0.0197 | 71.763 | $2.3233810^{-64}$ |
| $\alpha$ | 1/6 | - | - | - |
| $\delta$ | 0.095 | 0.0011 | 85.930 | $1.803610^{-69}$ |
| $\gamma$ | 1/21 | - | - | - |

**Figure 2:**
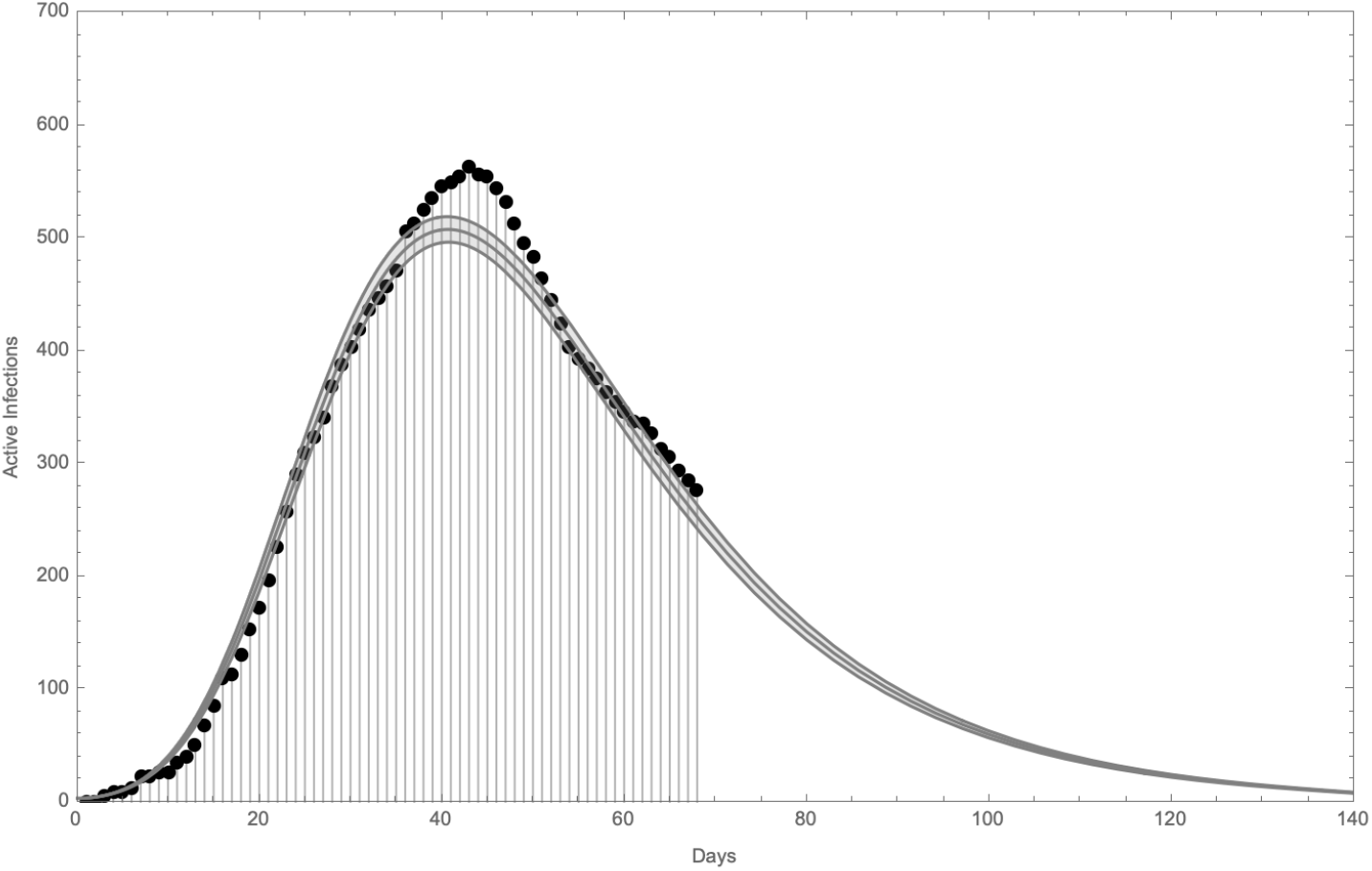
Best fit SEICR compartment model for Costa Rica. The 99% intervals are given with adjusted *R*^2^ = 0.99

The fitted value for transmission rate *β* = 1.416. The high value of confinement rate *δ* = 0.095, that is, roughly 1 person in 10 per day start confinement from the beginning of the epidemic. This is consistent with the early NPI measures applied by the government, but could be also the added effect of capturing quickly first cases and contact tracing, rendering an effective quick reduction of available susceptible.

### 5.2 Second Wave Projections

To explore how a second wave would unfold, we adjusted the data to include the last days where the number of cases started rising again (until May 14). The goal was to obtain an approximation of *ξ* which is the return to susceptible rate, simulating the relaxation of social distancing measures. With the available data was difficult to estimate, but we found a rough approximation of *ξ* ≈ 0.003, which corresponds to 3% relaxation of *δ*. From there we simulated a situation where the social distancing measures are reinstated after 22 days and 30 days as seen in figure 3, with some intervals of ±0.001. To achieve this we used the following definiton of *ψ*(*t*)

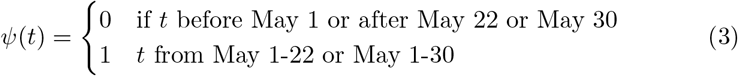

**Figure 3:**
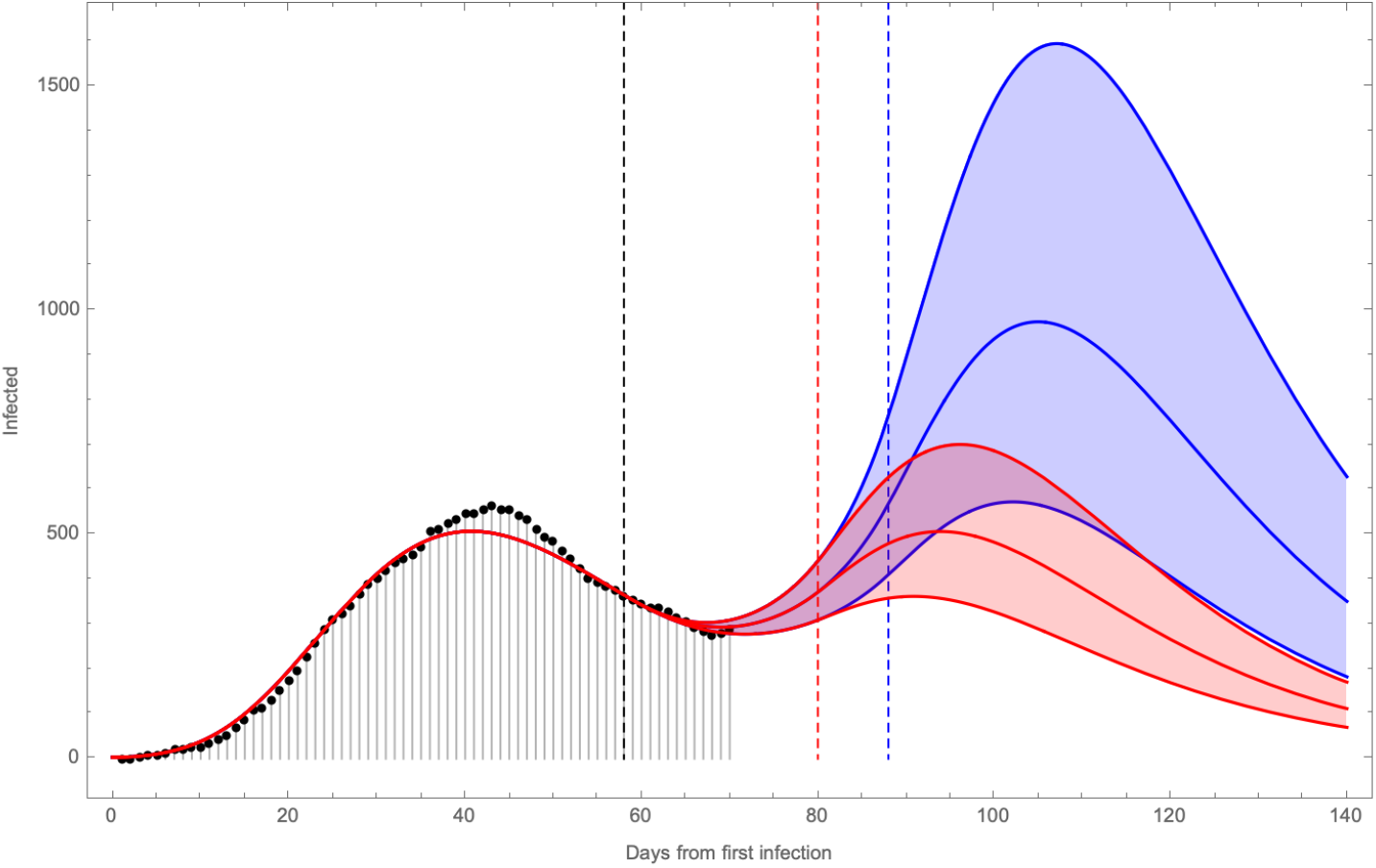
Second wave scenarios. The black dashed line corresponds to May 1*^st^*, where some restrictions where lifted. The red dashed line corresponds to hypothetically re-instating restrictions after 22 days, and the blue after 30 days. The upper and lower bounds are given by varying *ξ* by ±0.001

Which simulates the relaxation for a fixed period of time. At the time of writing this paper 15 days of relaxation have passed.

## 6 Discussion and Conclusion

### 6.1 Possible Interpretation of the Model

- The short lived first wave imposed difficulties when working with a simple SIR or SEIR models for Costa Rica. A sub-exponential increase of the epidemic in the first phase, made these models not suitable to explain COVID dynamics in Costa Rica.
- This model did have a good fit with the observed sub-exponential growth
- The sub-exponential increase, could be explained if we assume that the health ministry was quick in caching the very first cases and then, thru contact tracing, all possible exposed individuals from those very first cases.
- The movement of individuals from susceptible to confinement in the SE-ICR model, allows for the change dynamic behaviour without altering transmission rate, in much the same way as in [9].
- The inclusion of a Confined compartment allowed for adjusting the real potential number of susceptible people, as opposed to assuming that the entire Costa Rica population was susceptible. We could interpret that the fast isolation of first cases and their contacts, implicitly resulted in a fast reduction of the potential susceptible population (parameter *δ* in the SEICR model).
- We assume that *δ* captures NPIs in such a way, that people reduce contact and effectively move away from the susceptible population. It is possible that the population, after the emergency was declared and schools were closed (March 16*^th^*), kept contact cells to a minimum (only immediate family), effectively reducing the susceptible population (Google mobility data shows that behaviour)
- If fast catching of infected has occurred, then the transmission rate *β* = 1.416 reflects only infections within the contact tracing data, and not necessarily COVID transmission rate.
- The model did have a good fit using parameter values for *α* and *γ* that are infromal estimates for Costa Rica.
- Second wave predictions are highly sensitive to *ξ*. As shown in figure **??**, depending on when when restrictions are imposed, the second wave could have a peak of 250 cases, to as much as 1600. further validation of *delta* and *ξ* is needed.
- The SEIRCS model can be used constantly readjusting for new daily data and to asses possible trajectories based on current NPI measures
- As seen in figure 3, and somewhat expected, the reinstating of NPIs has a delayed effect on the control of the epidemic. This has to be taken into consideration, when planning NPI.
- With the SEICRS model cyclic strategies similar to the one proposed in [7] (10-4 strategy) could be studied. However, validation of parameters *δ* and *ξ* is required for this type of prediction

### 6.2 Model Limitations

- The model assumes that the data in Costa Rica captures all infected cases, but COVID incidence could be much higher, since there is no population level sample, or information on asymptomatic cases.
- There is no data on asymptomatic
- We have no data available to validate parameter *δ* and *ξ*. The model is highly sensitive to small variations of these parameters.
- As recognized by the government the pandemic in Costa Rica has shown an “atypical” behaviour [4], that is, there has been sub-exponential increase of cases and low mortality. This could mean that there are a large amount of confounding factors at population, genetic, cultural and molecular levels.
- No information is available publicly on the origin of new cases, this makes it difficult to filter data to show the effect of NPI.
- The Health ministry has indicated (without showing detail data) that there are no community infections.
- It is assumed that the government NPIs and population response are effectively having a reduction of “available” susceptible, by removing part of the population from freely mixing in the population.

## Data Availability

Data for active COVID-19 cases used for the model was obtained directly from the Ministry of Health website in Costa Rica.

https://www.ministeriodesalud.go.cr/index.php/centro-de-prensa/noticias/741-noticias-2020/1532-lineamientos-nacionales-para-la-vigilancia-de-la-infeccion-por-coronavirus-2019-ncov

